# Endophenotype-Informed Polygenic Risk Stratification for Primary Open-Angle Glaucoma in individuals of African Ancestry

**DOI:** 10.1101/2025.09.29.25336758

**Authors:** Aude Benigne Ikuzwe Sindikubwabo, Lannawill Caruth, Yan Zhu, Yuki Bradford, Colleen Moorse, Mina Halimitabrizi, Leila Ghaffari, Rebecca Salowe, Roy Lee, Anurag Verma, Penn Medicine BioBank, Regeneron Genetic Center, Marijana Vujković, Ahmara Gibbons Ross, Joan O’Brien, Shefali Setia-Verma

**Affiliations:** Department of Pathology and Laboratory Medicine, University of Pennsylvania, Philadelphia, PA 19104, USA; Center for Genetics of Complex Disease, Department of Ophthalmology, University of Pennsylvania Philadelphia, PA 19104, USA; Department of Medicine, Division of Translational Medicine and Human Genetics, University of Pennsylvania-Perelman School of Medicine, Philadelphia, PA 19104, USA; Department of Medicine, University of Pennsylvania, Philadelphia, PA 19104, USA; Department of Ophthalmology, University of Pennsylvania, Philadelphia, PA 19104, USA; Institute for Biomedical Informatics, University of Pennsylvania - Perelman School of Medicine, Philadelphia, PA 19104, USA; Division of Informatics, Department of Biostatistics, Epidemiology, and Informatics, University of Pennsylvania, Philadelphia, PA 19104

**Keywords:** Glaucoma, Primary Open-Angle Glaucoma, Genome-wide association analyses, glaucoma-associated traits, endophenotypes, mediation analyses, risk prediction, polygenic risk score, endophenotype informed polygenic risk score

## Abstract

Primary open-angle glaucoma (POAG) is a leading cause of irreversible blindness, yet up to half of cases remain undiagnosed, limiting opportunities for early intervention. Genetic risk stratification may improve early detection, particularly in people of African ancestry, who experience a disproportionate burden of POAG but remain underrepresented in genetic studies. Prior glaucoma polygenic risk score (PGS) approaches have incorporated POAG and other related ocular traits, including intraocular pressure (IOP) and cup-to-disc ratio (CDR) to improve prediction. However, most evaluations have emphasized aggregate discrimination, while less is known about whether endophenotype-specific genetic information can improve clinical interpretability or identify distinct individuals at the extremes of risk. Here, we developed endophenotype-informed PGS models that preserve genetic information related to IOP and CDR as distinct components of risk. These scores were evaluated in an independent testing cohort from the Primary Open-Angle African Ancestry Glaucoma Genetics (POAAGG) study (n=271), validated in the Penn Medicine Biobank (PMBB) cohort (n=1,662), and assessed for clinical relevance in a longitudinal cohort of POAAGG glaucoma suspects. We also performed manual chart reviews of PMBB participants in the highest and lowest extremes (5th percentiles) of PGS distribution to evaluate whether endophenotype-informed scores identified clinically meaningful patterns of risk.

Endophenotype-informed PGS provide pathway-level attribution not available from a single aggregate POAG PGS score. Although models incorporating measured IOP and CDR as covariates achieved similar overall discrimination, they did not prioritize the same individuals at the extremes of predicted risk. In the longitudinal suspect cohort, the baseline IOP-informed score stratified individuals who later converted to POAG from those who remained stable. Manual chart review further supported the clinical interpretability: the CDR-informed score identified structural susceptibility to glaucomatous damage beyond EHR-based classification, while the IOP-informed score captured genetic susceptibility to pressure-related disease. Together, these findings show that endophenotype informed polygenic modeling can move glaucoma genetic risk prediction beyond aggregate discrimination toward a more clinically interpretable framework for risk stratification. In African ancestry populations, this approach may help identify biologically meaningful heterogeneity, support conversion-risk prediction, and prioritize individuals who could benefit from earlier surveillance.

## INTRODUCTION

Primary open-angle glaucoma (POAG) is a chronic eye disease characterized by progressive damage of the optic nerve and corresponding loss of visual fields.^1^ POAG is the most prevalent glaucoma subtype, affecting 68 million individuals worldwide, with a global prevalence of 2.2%.^2,3^This heritable disease disproportionally affects individuals of African ancestry, who are four to five times more likely to develop the disease compared to individuals of European ancestry.^4^ Moreover, approximately half of individuals with POAG remain undiagnosed until significant vision loss has already occurred.^4^ POAG’s prevalence is projected to increase by 79.8% globally by 2040, with individuals of African ancestry contributing a disproportionately high 130.8% rise in cases.^5^ These trends underscore the urgent need for novel screening tools to identify high-risk individuals before irreversible optic nerve damage and vision loss occur.

Beyond ancestry, risk factors for POAG include older age, positive family history and a substantial inherited genetic component, with heritability estimates ranging from 17 to 81%.^6^ Several heritable ocular traits, often referred to as endophenotypes are associated with POAG. Endophenotypes are continuous, clinical measures that contribute to disease susceptibility and are routinely used in the diagnosis and monitoring of POAG.^4,6^ Although these traits lie on the causal pathway to POAG (e.g., elevated intraocular pressure [IOP], cup-to-disc ratio [CDR], and retinal nerve fiber layer [RNFL]), they are not independently causal; instead, they are informative markers of disease risk and progression.^7^ These traits also demonstrate significant heritability, particularly IOP (0.43 [0.38–0.48]), CDR (0.56 [0.44–0.68]), and RNFL thickness (0.73 [0.42–0.91]).^6^ Notably, genome-wide association studies (GWAS) have shown that up to 72% of loci significantly associated with POAG also show associations with IOP,^8^ and approximately 40% of loci linked to CDR are also associated with POAG.^9^ This pattern of shared genetic architecture highlights the clinical relevance of these endophenotypes in understanding the underlying biology of POAG.

Such cross-phenotype genetic associations are common in complex diseases such as POAG, where genetic variants may influence both disease status and clinically relevant endophenotypes.^10^ However, it is unclear whether these associations reflect independent genetic effects on multiple traits (biological pleiotropy) or effects transmitted through an intermediate phenotype (mediated pleiotropy).^10^This distinction has important implications for clinical translation. While inherited genetic risk is not modifiable, the intermediate traits through which genetic risk is expressed may be clinically measurable and actionable. Identifying genetic effects that operate through these intermediate traits could improve both the predictive performance and interpretability of genetic risk models, with potential integration into targeted screening and prevention strategies.

Genome-wide association studies have identified numerous variants associated with POAG and its ocular endophenotypes,^11,8,12,13,14,4,9^ including IOP and CDR, providing the foundation for polygenic risk scores (PGS) that estimate inherited disease susceptibility. However, these polygenic risk scores typicaly aggregate genetic effects into a single risk estimate. In some prediction models, these scores are further combined with clinically measured endophenotypes, such as IOP or CDR, by including those traits as covariates. s. These approaches assumes the genetic and endophenotype contributions to POAG risk are additive, and does not explicitly model the possibility that a variant’s contribution to disease risk depends on an individual’s endophenotype value, for example, through pathway-specific mediation of genetic effects via IOP or CDR. Critically, this aggregation also obscures which loci act through a given endophenotype, which act independently of it, and which act through unmeasured pathways, information that is not recoverable once genetic effects are summarized into a single score. To address this limitation, we leveraged mediation-aware GWAS to decompose variant effects into disease-associated components that are mediated through clinically measured endophenotypes and components that are independent of them. This framework in turn enabled construction of endophenotype - informed PGS that integrates genetic susceptibility with an individuals’ measured IOP and CDR value, preserving pathway-specific attribution within the risk model..^4,15^ By linking genetic risk prediction to measurable clinical pathways, this approach may produce scores that are not only predictive, but also more biologically interpretable and clinically informative. Although, this framework has demonstrated promising results for liver disease,^15^ but its utility for understanding the genetic architecture and risk pathways of POAG remains unexplored.

We used the Primary Open-Angle African Ancestry Glaucoma Genetics (POAAGG) cohort and the Penn Medicine Biobank (PMBB) to systematically disentangle the genetic architecture underlying POAG and its key endophenotypes and to evaluate genetically informed approaches for POAG risk stratification in individuals of African ancestry. We generated and compared multiple sources of genetic risk information, including genome-wide POAG association estimates, multi-trait association estimates integrating POAG and related endophenotypes, selected SNP sets derived from these GWAS, and endophenotype-informed estimates from a mediation framework. We then constructed PGS models from these complementary genetic architectures and evaluated their performance alone and in combination with clinically measured endophenotypes, including IOP and CDR. Finally, we assessed model performance across testing, an independent testing, external validation, and longitudinal glaucoma suspect cohorts, with manual chart reviews to evaluate clinical interpretability. By integrating genetic risk modeling with clinically measured ocular traits, this study evaluates whether endophenotype-informed approaches can support more interpretable POAG risk stratification and precision screening in a population disproportionately affected by the disease.

## METHODS

### Study Datasets

#### POAAGG Study

The POAAGG study cohort consists of self-identified Blacks (African American, African descent, or African Caribbean), aged 35 years or older, recruited from the Philadelphia region between 2010 and 2019. Subjects were enrolled during regularly scheduled appointments at the Ophthalmology Department at the University of Pennsylvania and other sites.^16^ At the time of enrollment, each subject was classified as a POAG case, suspect, or control by a glaucoma specialist or ophthalmologist based on previously published criteria and provided a genomic DNA sample.^17^ All subjects provided written informed consent for their participation in the study. The POAAGG study was approved by the University of Pennsylvania Institutional Review Board under protocol #812036 and followed the tenets of the Declaration of Helsinki.

During the study visit, subjects underwent an onsite interview and examination, which included the collection of baseline endophenotypes. These measures included IOP, CDR, retinal nerve fiber layer (RNFL) thickness, visual acuity (VA), central corneal thickness (CCT), visual field mean deviation (MD), and pattern standard deviation (PSD). For subjects with IOP-lowering medication or a history of glaucoma surgery *prior* to the IOP_baseline measurement, pre-treatment IOP was calculated by dividing IOP_baseline by 0.7. This particular estimate was based on prior literature demonstrating reduction of IOP levels by 30% following IOP-lowering medications^18^ and 32% following trabeculotomy.^19^ Maximum intraocular pressure (IOP_max) was defined as the highest IOP value from all IOP measurements, from baseline to most recent visit, for each subject. All ocular characteristics were stored in Research Electronic Data Capture (Table 1). ^20,21^ For each of the endophenotypes, the value from the more severely affected eye was used in this study.

**Table 1:** Description of endophenotypes, corresponding sample sizes, and demographics for POAAGG cohort.

|  |  |  |  |  |  |  |  |  |
| --- | --- | --- | --- | --- | --- | --- | --- | --- |
| **Endo phenotypes** | **Total N** | **Cases (N)** | **Controls (N)** | **Phenotype Mean ± SD (Cases)** | **Phenotype Mean ± SD (Controls)** | **Mean Age ± SD** | **Females (N)** | **Males (N)** |
| CDR | 5181 | 2285 | 2810 | 0.76 ± 0.16 | 0.33 ± 0.13 | 65.5 ± 12.01 | 3054 | 1691 |
| IOP_max | 5499 | 2173 | 3223 | 18.05 ± 6.43 | 15.62 ± 2.89 | 65.0 ± 12.07 | 3291 | 1750 |
| IOP_baseline | 5565 | 2223 | 3239 | 21.71 ± 7.71 | 15.73 ± 3.03 | 65.1 ± 12.07 | 3328 | 1776 |
| PSD | 2025 | 1909 | 105 | 6.43 ± 3.65 | 4.14 ± 3.39 | 69.2 ± 11.02 | 1134 | 779 |
| VA | 5397 | 2083 | 3246 | 0.23 ± 0.42 | 0.18 ± 0.38 | 64.7 ± 12.06 | 3271 | 1696 |
| CCT | 2684 | 2219 | 430 | 529.14 ± 38.12 | 541.92 ± 41.15 | 68.6 ± 11.45 | 1511 | 978 |
| MD | 2030 | 1914 | 105 | -10.39 ± 9.48 | -4.82 ± 6.49 | 69.1 ± 11.02 | 1134 | 785 |
| RNFL | 2079 | 1866 | 208 | 68.73 ± 14.09 | 83.42 ± 13.05 | 68.3 ± 11.27 | 1175 | 780 |
| CCT=Central Corneal Thickness, CDR=Cup-to-disc ratio, IOP_baseline=Intraocular Pressure at baseline adjusted for either medication or surgery, IOP_max= Maximum Intraocular Pressure, MD = Mean Deviation, PSD = Pattern Standard Deviation, RNFL = Retinal Nerve Fiver Layer Thickness, VA = Visual Acuity, logMAR = Logarithm of the Minimum Angle of Resolution |  |  |  |  |  |  |  |  |

Details of the study design are shown in Figure 1. POAAGG subjects were divided into the following cohorts for this study:

- **POAG-Related Endophenotypes cohort**: This cohort was used to perform both the GWAS of POAG-related endophenotypes and the endophenotype-adjusted GWAS. A total of 6,229 subjects (2,696 cases and 3,533 controls) were included. Inclusion criteria included defined case or control status and available baseline measurements for at least one endophenotype.
- **PGS testing cohort:** This cohort was used to evaluate the performance of endophenotype-informed PGS. This cohort consisted of 271 POAAGG subjects, including 128 cases and 143 controls, who were held out from the GWAS analyses. To confirm accurate case–control classification, phenotypic data for each subject were carefully reviewed. Of the 271 subjects, 196 were selected as clean cases and healthy controls based on strict phenotypic criteria. Cases were required to meet the following thresholds: IOP ≥ 22.1 mmHg, CDR ≥ 0.7, and RNFL ≤ 80 μm. Controls were required to have normal phenotypes, defined as IOP between 10–16 mmHg, CDR ≤ 0.5, and RNFL ≥ 82 μm. An additional 75 subjects were included from recent visits within the same healthcare system.
- **Suspects cohort:** This cohort of suspects from the POAAGG study was used to evaluate how well the models distinguished between “converters” (converted from suspect to case over time) and “non-converters” non-converters (remained table throughout the available ophthalmic follow-up, with no evidence of progression to POAG).^22^ The cohort included 248 POAAGG suspects with longitudinal follow-up data, including 28 converters and 220 non-converters.^22^ Endophenotype-informed PGS were calculated for these subjects. These individuals were not included in the PGS model performance assessment.

**Figure 1.**
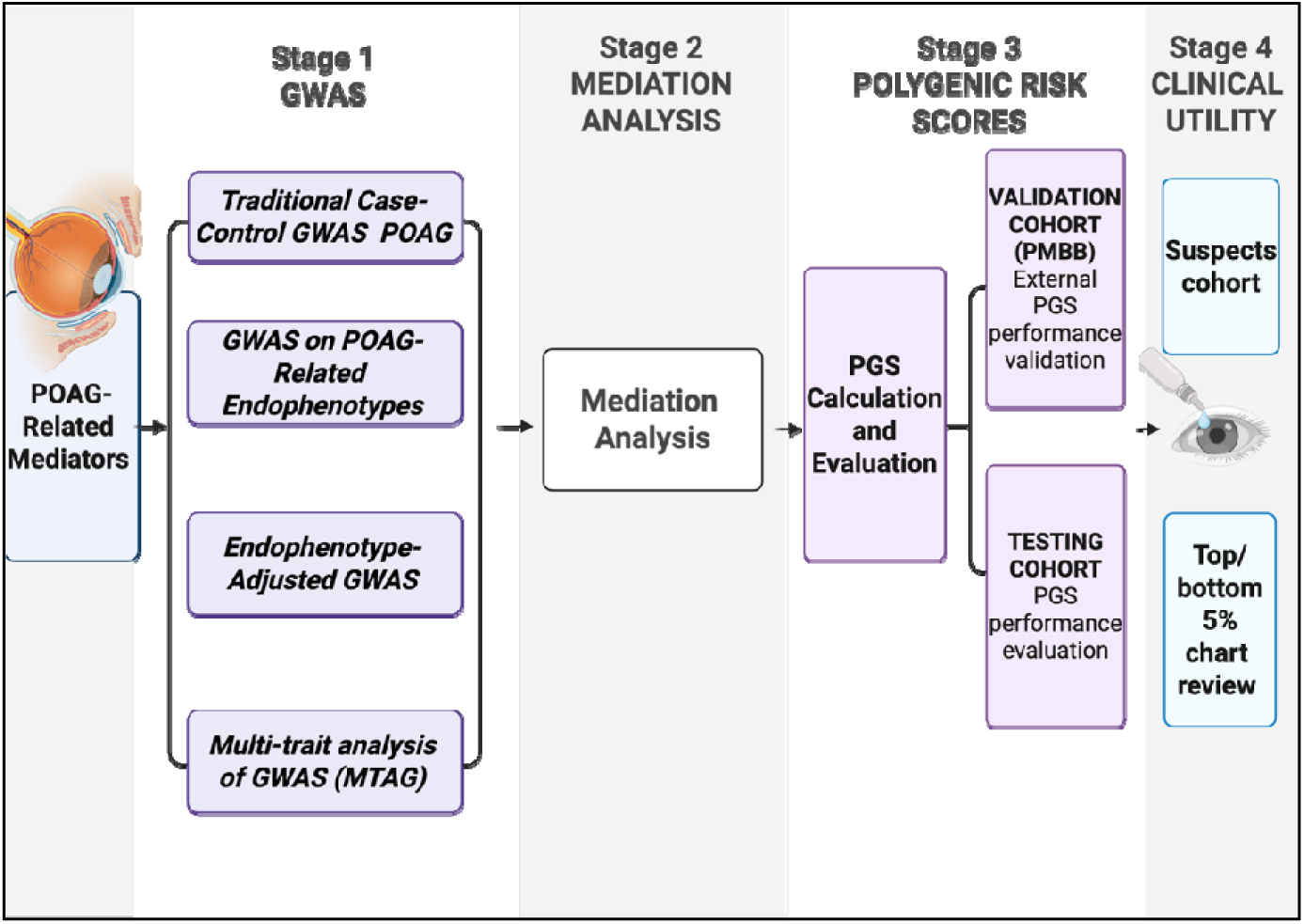
Flowchart illustrating the overall study design and analytical workflow.

#### Penn Medicine Biobank (PMBB)

The Penn Medicine Biobank (PMBB) was used as an independent PGS validation cohort. PMBB is a large health system-based biorepository embedded within the University of Pennsylvania.^23^ We identified participants of African ancestry, aged ≥35 years, using the same inclusion/exclusion criteria as the POAAGG study.^16^ POAG cases were identified using International Classification of Diseases (ICD-10) code H40.11. Controls were defined as individuals without a POAG diagnosis, who had at least one ophthalmology visit and no history of IOP-lowering treatment, to minimize misclassification due to undiagnosed disease. In addition, controls with potential confounding ocular conditions were excluded; the ICD codes used to define these exclusion criteria are provided in Supplementary Table S1.

From this population, we further selected participants with both baseline IOP and CDR measurements available. Pre-treatment IOP was calculated for participants with IOP-lowering medication or a history of glaucoma surgery, including laser interventions and incisional glaucoma surgeries using the same methodology as for the POAAGG cohort. To ensure independence from the discovery cohort, individuals overlapping between the PMBB and POAAGG cohorts were excluded. ^16^The final PMBB analytic dataset included 1,485 controls and 177 cases. This cohort was used to evaluate the performance of endophenotype-informed PGS as well as other different polygenic risk score models. The analyses in this cohort were restricted to IOP and CDR to align with the mediation-informed PGSs generated in the discovery cohort. Details of the PMBB cohort are presented in Table 2.

**Table 2:** Description of endophenotypes, corresponding sample sizes, and demographics for African PMBB validation cohort.

| Endo phenotypes | Total N | Cases (N) | Controls (N) | Phenotype Mean ± SD (Cases) | Phenotype Mean ± SD (Controls) | Mean Age ± SD | Females (N) | Males (N) |
| --- | --- | --- | --- | --- | --- | --- | --- | --- |
| CDR | 1662 | 177 | 1485 | 0.62 ± 0.22 | 0.37 ± 0.17 | 59.0 ± 11.38 | 1059 | 603 |
| IOP_baseline | 1662 | 177 | 1485 | 20.99 ± 8.36 | 15.77 ± 3.24 | 59.0 ± 11.38 | 1059 | 603 |

CDR=cup-to-disc ratio, IOP\_baseline=Intraocular Pressure at baseline adjusted for either medication or surgery

### Overview of Analysis

We developed an endophenotype-informed genetic risk modelling framework to evaluate whether incorporating clinically measurable POAG-related traits could improve risk stratification and biological interpretability. Traditional GWAS estimates the total association between genetic variants and disease status (i.e., POAG) but does not distinguish whether these associations act directly on disease risk or through intermediate traits. By incorporating IOP and CDR, two well-characterized POAG endophenotypes, our approach enables decomposition of total genetic effects into (1) direct effects on POAG and (2) indirect effects transmitted through these measurable traits.

Our analysis was conducted in 4 main stages (Figure 1): *Stage 1*: Genome-Wide Association Studies. This stage included a traditional case-control GWAS of POAG, GWAS of POAG-related endophenotypes, an endophenotype-adjusted GWAS for POAG as outcome, and a multi-trait analysis of GWAS (MTAG). *Stage 2* : Mediation analysis. This stage included mediation analyses to partition SNP effects into direct and endophenotype-mediated components, followed by proportion mediated calculations, *Stage 3*: Polygenic Risk Score Construction and Evaluation. This stage included construction of an endophenotype-informed PGS, as well as construction of other previously established PGS models followed by evaluation of predictive performance across testing and validation cohorts. *Stage 4*: Evaluation of Clinical Utility. This stage assessed discrimination between converters and non-converters in the POAAGG glaucoma suspect cohort, and manual chart reviews of PMBB participants with discordant disease status and PGS-predicted risk.

Each of these analyses are described in detail in the sections below. Of note, while POAG is influenced by multiple ocular traits, stages 2, 3 and 4 were restricted to IOP and CDR, two of the most clinically established and routinely measured POAG-related traits. IOP is also directly modifiable through existing IOP-lowering therapies, making this framework particularly relevant for translation into risk-based screening and prevention strategies.

### Genotype Data: Quality Control and Imputation

The POAAGG dataset was imputed using the TOPMed reference panel R2. ^4,24^ Post-imputation quality control was applied, excluding SNPs with an average imputation R^2^<0.3, minor allele frequency (MAF) <1%, and/or significant deviation from Hardy–Weinberg equilibrium (p < 1 × 10□□). Principal component analysis (PCA) was conducted with the EIGENSOFT package, projecting the PCs onto the 1000 Genomes dataset.^4,25,26^ Similarly, genotype data from the PMBB were obtained from the 2024 (V3) data release. Genotypes were imputed using the TOPMed R3 (GRCh38) reference panel. Post-imputation quality control excluded variants with an imputation quality score (R²) < 0.2, MAF < 1%, and significant deviation from Hardy–Weinberg equilibrium (p < 1 × 10^□6^). PCA was conducted using EIGENSOFT to account for population structure and infer genetically informed ancestry.

### Stage 1: Genome-Wide Association Studies

#### Traditional Case-Control GWAS of POAG

First, to contextualize and evaluate the findings from our mediation framework, we used our previously published POAG GWAS^4^ as a baseline reference. This study was a mega-analysis conducted in three datasets that prioritized patients of African ancestry, including the POAAGG cohort, the African Descent and Glaucoma Evaluation Study (ADAGES), and the Genetics of Glaucoma in People of African Descent (GGLAD) consortium^4^, totaling 11,275 subjects of African ancestry (6,003 cases, 5,272 controls, excluding POAAGG testing cohort samples). The mega-analysis as previously described identified 46 genome-wide significant loci associated with POAG using a traditional case-control GWAS approach, without adjusting for endophenotypes.^4^

#### GWAS on POAG-Related Endophenotypes

We performed a GWAS using a mixed linear regression model for each of the eight POAG-related endophenotypes (also referred to as mediators), as described in Eq. (1), using SAIGE version 1.2.0^27^. For each participant, the baseline clinical measurement was used when available. Because these measurements were obtained as part of routine clinical care, baseline values do not necessarily represent measurements collected before POAG onset and, in affected individuals, may already reflect early disease-related changes. SNP-level association tests were conducted to quantify the genetic influence on each mediator. The baseline values were adjusted if patients were on IOP lowering treatment or had surgery in the past, as described above.^18,19^Age, sex and the first five principal components (PCs) were used as covariates. SAIGE applies a mixed linear regression model while also accounting for sample relatedness. For continuous traits such as these endophenotypes, SAIGE also performs inverse normal transformations to ensure a normal distribution.^27^

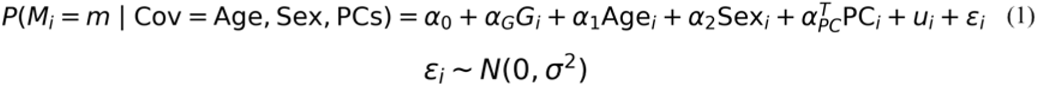

Association of Endophenotypes/Mediator with SNPs

Where:

- □ denotes an individual

-*M*□ denotes each of the mediator

- *G*□ ∈ {0, 1, 2} is the genotype dosage at the variant of interest,

- *á_G_* quantifies the effect of genotype on the mediator,

- *PC*□ is the vector of top genetic PCs

- *u*□ ∼ N(0, τK) is a random effect accounting for sample relatedness, with K denoting the genetic relationship matrix (GRM).

#### Endophenotype-Adjusted GWAS

Next, to evaluate the effect of genetic variants on POAG, we conducted a series of POAG case-control regression GWAS analyses, adjusting for each of POAG’s endophenotypes as shown in Table 1 along with other baseline covariates (age, sex and PCs). This approach allows for estimation of SNP effects on POAG conditional on the mediator, as described in Eq. (2).^27^ We compared the SNP effect sizes from each mediated GWAS to those from our traditional case-control POAG GWAS (no mediator adjustment).^4^ This comparison distinguished direct genetic effects from those potentially operating through the mediator. Age, sex and the first five PCs were used as covariates. The binary outcome Y ∈ {0,1} was modeled using a logistic mixed model, as implemented in SAIGE:

Association of Outcome (POAG) with SNPs adjusted by mediator/endophenotypes:

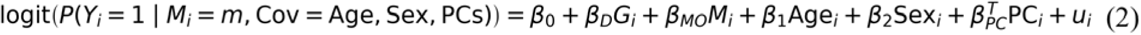

Where:

- □ denotes an individual,

- β*_D_* represents the direct effect of genotype on the outcome, adjusting for the mediator,

- β*_MO_* captures the effect of the mediator on the outcome,

- *PC* is the vector of top genetic PC,

- u∼ N(0, τK) is a random effect accounting for sample relatedness, with K denoting the genetic relationship matrix (GRM).

#### Multi-trait analysis of GWAS (MTAG)

We performed a Multi-Trait Analysis of GWAS (MTAG; v1.0.8) ^28^using GWAS summary statistics from the traditional case-control POAG GWAS, baseline IOP GWAS, and CDR GWAS described above. MTAG jointly analyzes genetically correlated traits to improve SNP effect size estimation while accounting for sample overlap.

### Stage 2: Mediation analysis

#### Compiling Variants of Interest

The mediation analysis framework requires genetic variants to be significantly associated with the outcome of interest^15^. Here, to compile the list of POAG-associated variants, we curated all genome-wide significant loci reported across five previously published multi-ancestry POAG GWAS studies. ^4, 8,11,12,13^ The combined list all these studies are in Supplementary Table S2. Because the POAAGG genotype data were aligned to the GRCh38 (hg38) reference genome, variants reported in earlier genome builds were lifted over to hg38 prior to analysis. The resulting variant set was then subjected to quality control, retaining only SNPs that were successfully mapped and present in the POAAGG cohort. After these filtering steps, 568 SNPs remained for downstream mediation analyses. The final set of 568 quality-controlled SNPs are provided in Supplementary Table S3

#### Estimation of Direct vs Indirect Genetic Effects for IOP and CDR

To estimate the extent to which an endophenotype mediates the relationship between each genetic variant and POAG, we conducted mediation analyses using four regression models per variant using the 568 SNPs described above. SNP to outcome association analyses (Models 1, 2, and 3) were performed using SAIGE^27^ as described above, whereas the mediator-to-outcome coefficient required for the indirect effect was estimated using PLINK v2.0.^29^

- Model 1 (total effect): POAG was regressed on the SNP and covariates using logistic regression, yielding the total effect of the SNP on POAG risk (*β_t_, SE*) on the log-odds scale.
- Model 2 (direct effect and mediator-on-outcome effect): POAG was regressed on the SNP, endophenotype, and covariates using logistic regression. This model yields: the direct effect of the SNP on POAG, independent of IOP or CDR (*β_D_, SE_D_*) on the log-odds scale. To estimate the effect of endophenotype on outcome, the same regression model was fitted using PLINK v2.0^29^ after adjusting for SNPs and covariates using logistic regression. This model estimates the effect of the endophenotype on POAG after adjustment for the SNP (*β_MO_, SE_MO_*) on the log-odds scale.
- Model 3 (SNP-on-endophenotype effect): IOP or CDR was regressed on the SNP and covariates using linear regression, yielding the effect of the SNP on IOP (*α_G_, SE_G_*) on the natural (continuous) scale of endophenotype.
- Indirect effect. The indirect effect (IE) of the SNP on POAG transmitted through endophenotype was estimated using the product-of-coefficients method.^15^

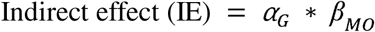 This approach is appropriate when the mediator is continuous and the outcome is binary, as it operates on compatible scales: *α_G_* quantifies the per-allele change in endophenotype in its natural units, and *β_MO_* quantifies the change in log-odds of POAG per unit of endophenotype. Their product gives the indirect effect on the log-odds scale. The standard error of IE was estimated using the Sobel formula.^30^

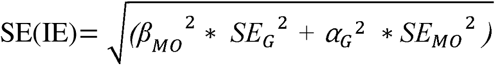 Statistical significance of the indirect effect was evaluated as a two-sided z-test: Z = IE / SE(IE), with the corresponding p-value derived from the standard normal distribution.

#### Proportion Mediated

Because POAG is a binary outcome modelled on the log-odds scale, the proportion of the total genetic effect mediated through IOP was quantified using the formula of VanderWeele and Vansteelandt^31^ for binary outcomes, which accounts for the non-collapsibility of the odds ratio.

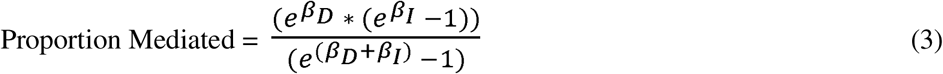

Where:

*β_D_* represents the direct effect of genotype on the outcome, adjusting for the mediator

*β_l_* is the indirect effect, both on the log-odds scale.

#### Significance of Mediation and Classification for IOP and CDR

To formally test whether the direct and total genetic effects differed significantly and therefore whether meaningful mediation was present, we computed the Z-difference statistic^32^:

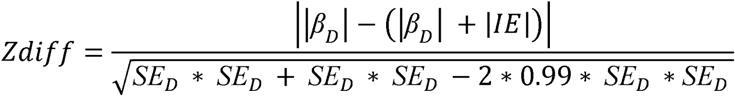

This assumes approximate independence between the total and direct effect estimates, which is standard when working with GWAS summary statistics. The corresponding two-sided p-value (P_diff) was obtained from the standard normal distribution, with P_diff < 0.05 considered statistically significant.

Variants were classified following the criteria, as described and implemented in Vujkovic et al. ^15^

- **Full mediation**: proportion mediated > 70% and P_diff < 0.05
- **Partial mediation:** proportion mediated ≤ 70% but P_diff < 0.05
- **No significant mediation:** P_diff ≥ 0.05, regardless of the estimated proportion
- **Inconsistent (Non interpretable**): *β_D_*and IE have opposite signs, indicating the mediator suppresses rather than transmits the genetic effect; these variants were not interpreted as mediating

### Stage 3: Polygenic Risk Score Construction and Evaluation

#### Previously Established Polygenic Risk Scores

##### 568-SNP PGS

Using the same set of 568 SNPs described above, effect size estimates of those variants were obtained from an African ancestry-specific GWAS described above^4^ as well as MTAG summary statistics described above to ensure appropriate ancestry-matched weighting. PGS scores were calculated using PLINK v2.0^29^ and subsequently normalized prior to downstream analyses.

##### Genome-wide PGS

To construct genome-wide polygenic risk scores, we applied PRS-CSx^33^, a Bayesian polygenic prediction method that estimates posterior SNP effect sizes while accounting for linkage disequilibrium (LD), separately to (1) the summary statistics from the traditional case-control POAG GWAS alone, and (2) the MTAG-derived summary statistics. The resulting posterior effect size estimates from each analysis were used to calculate two genome-wide PGS: a POAG-only PRS-CSx PGS and an MTAG-derived PRS-CSx PGS.

#### Endophenotype-Informed PGS for IOP and CDR

An endophenotype-informed PGS was constructed separately for IOP and CDR endophenotypes. This approach models the hypothetical (or “counterfactual”) outcome^34,35,36^ that would occur under different levels of the mediator, enabling decomposition of total SNP effects into components mediated by endophenotypes versus those acting directly on disease. Briefly, this framework extends standard regression-based mediation to the high-dimensional, genome-wide setting by using effect estimates from GWAS of both the mediator and the outcome, along with joint modeling of covariates. Our implementation leverages these estimates to isolate the direct (non-mediated) SNP effects, which form the basis of the endophenotype-informed PGS.^15^

Mediator-Informed PGS:

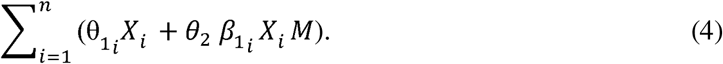

Where:

*θ*_1_ is the direct effect from SNP to POAG,

*β*_l_ is the direct effect from SNP to mediator

θ_2_ is the indirect effect of SNP to POAG through the mediator

M is the value of the mediator.

#### Evaluation of all Polygenic Risk Scores

##### Evaluation of Previously Established PGSs

To benchmark the proposed endophenotype-informed PGS against existing polygenic risk prediction approaches, we conducted a series of sensitivity analyses in the POAAGG testing cohort and PMBB validation cohort using logistic regression models. Nine models were evaluated: (1) baseline covariates (age, sex, and PC1–PC4); (2) baseline covariates plus measured intraocular pressure (IOP); and (3) baseline covariates plus measured cup-to-disc ratio (CDR). (4) the genome-wide PRS-CSx PGS derived from the traditional case-control POAG GWAS alone; (5) the genome-wide PRS-CSx PGS derived from the MTAG-derived summary statistics described above; (6) 568-SNP POAG PGS constructed from previously reported POAG-associated variants weighted by traditional case-control POAG GWAS ;(7) a 568-SNP MTAG PGS constructed using the same variant set but weighted by MTAG-derived effect estimates, (8) the 568-SNP POAG PGS combined with measured IOP; and (9) the 568-SNP POAG PGS combined with measured CDR. Predictive performance was evaluated using the area under the receiver operating characteristic curve (AUC–ROC), which measures the ability of the model to distinguish between cases and controls. Additional metrics included balanced accuracy, precision–recall area under the curve (PR-AUC), sensitivity, specificity, and Brier score, which together assess classification performance, discrimination of true cases and controls, and calibration of predicted probabilities.

##### Evaluation of Endophenotype-informed PGS

The endophenotype-informed PGS was evaluated using the same set of 568 QC-passed SNPs. The performance of the endophenotype-informed PGS was assessed using the PGS testing cohort from the POAAGG study and in validation cohort from PMBB. Model assessed were : an IOP-informed PGS (Model 10) and CDR-informed PGS (Model 11).Predictive performance was assessed using multiple metrics, including AUC-ROC, precision–recall area under the curve (PR-AUC), balanced accuracy, sensitivity, specificity, and Brier score to assess model performance as described above

### Stage 4: Evaluation of Clinical Utility

#### POAAGG Suspect Cohort

To further evaluate the ability of each endophenotype-informed PGS (IOP and CDR) model to differentiate individuals at risk of developing POAG, we applied both scores to the POAAGG suspect cohort, which includes converters and non-converters. For each model, we compared mean PGS values between non-converters and converters using an independent samples t-test, assessing whether suspects who converted to POAG carried a significantly higher polygenic burden than those who remained stable.

#### Chart Reviews

To further evaluate the clinical utility of the endophenotype-informed PGS, we examined PMBB participants at the extremes of the risk distribution. Participants were ranked according to their standardized PGS, and individuals within the top 5% (high risk) and bottom 5% (low risk) of the score distribution were identified. We then selected individuals whose clinical classification was discordant with their genetic risk prediction, which specifically included controls within the top 5% of the PGS distribution and POAG cases classified within the bottom 5%. These participants underwent detailed manual chart review to assess potential EHR misclassification and to identify evidence of subclinical glaucoma or other glaucoma-related abnormalities.

We reviewed the ophthalmic history for each participant, including history of glaucoma medications and surgeries, phenotypes (IOP, CDR, CCT, VA, RNFL, MD, and PSD), physician notes, and diagnosis notes at all available timepoints. Misclassifications were defined as the initial EHR-based classification of participant status (case, control, suspect) in PMBB by ICD codes was incorrect based on manual chart review of all available information. Conversion was defined as a change in participant status between their baseline and latest appointment (e.g., suspect to case). Among cases, we evaluated if participants met the criteria for normal-tension glaucoma (NTG) or ocular hypertension (OHT). NTG was defined as a glaucomatous optic neuropathy with characteristic optic nerve and visual field damage, despite IOP remaining within normal limits (i.e., untreated baseline IOP ≤ 21 mmHg and IOP_max ≤ 21 mmHg). ^37^ OHT was defined as consistently elevated IOP levels (untreated IOP between 24-32 mmHg in one eye, 21-32 mmHg in other eye), without detectable glaucomatous optic nerve damage or visual field loss.^38^

We also measured the rate of structural and functional progression among participants with longitudinal data. Structural progression was defined as the annual rate of change of global RNFL thickness (μm/year), and functional progression was defined as annual rate of change of MD from visual fields (dB/year). We calculated progression rates for each eye from linear mixed effects models with random intercepts and random slopes. We accounted for inter-eye correlation from the same participant and longitudinal correlation for the same eye over time. Best linear unbiased prediction was used to estimate individual slopes of RNFL thickness and MD for each eye.^39^

## RESULTS

### GWAS on POAG-Related Endophenotypes

A total of eight genome-wide significant loci (p-value < 5 × 10□□) were identified across the GWAS on each endophenotypes highlighting the distinct genetic architectures underlying these intermediate traits. The lead genome-wide significant locus for each endophenotype is summarized in Supplementary Table S4, the complete set of genome-wide significant association results is provided in Supplementary Table S5, and the corresponding Manhattan and Q–Q plots are shown in Supplementary Figure S1. We identified one significant locus associated with CCT (mapping to the nearest gene of *ZNF646)*, one with CDR (*LOC110120806)*, one with IOP_baseline (*LYPLAL1)*, one with IOP_max (*LOC105374760)*, one with PSD (*LENG8)*, and three with VA (*LOC107984525, SLC12A2,* and between *LOC124903246 / RPL35P9).* Of particular interest, *LYPLAL1* has previously been implicated in POAG and suggested as a potential drug target due to its protective effects on the disease.^40^ Consistent with a possible protective association, the variant that mapped on *LYPLAL1* in our IOP_baseline GWAS also showed a negative effect estimate (p-value: 4.10×10^-8^, Beta: -1.1065, SE: 0.201679), suggesting this variant is associated with lower IOP and may be protective against POAG.

### Genetic effects on POAG are mediated through IOP and CDR

We next sought to determine whether POAG-associated variants are mediated through IOP and CDR and, if so, to quantify the extent of this mediation. To begin, we used the same 568 SNPs that have been associated with POAG in prior studies, as described in the Methods. We then calculated the proportion mediated on all 568 SNPs (Supplementary Table S6 and S7). Because mediation analysis requires that the exposure (genetic variant) is associated with the endophenotype,^15^ we first tested the 568 candidate variants for association with each endophenotype (IOP and CDR). Using the nominal screening threshold of P<0.05, as described by Vujkovic et al.,^15^ 61 variants were associated with IOP and 72 with CDR. Each variant was then classified according to the mediation criteria, as described in the Methods.

Among the variants associated with IOP, 18 were classified as fully mediated and 18 as partially mediated (Figure 2). For CDR, 24 variants were classified as fully mediated and 15 as partially mediated (Figure 2). The remaining variants were classified as either showing no significant mediation or as non-interpretable. For visualization, variants that were either partially or fully mediated were annotated to their nearest genes. Because multiple variants could map to the same gene, only the variant with the strongest association with POAG (lowest p-value) was retained for each gene

**Figure 2:**
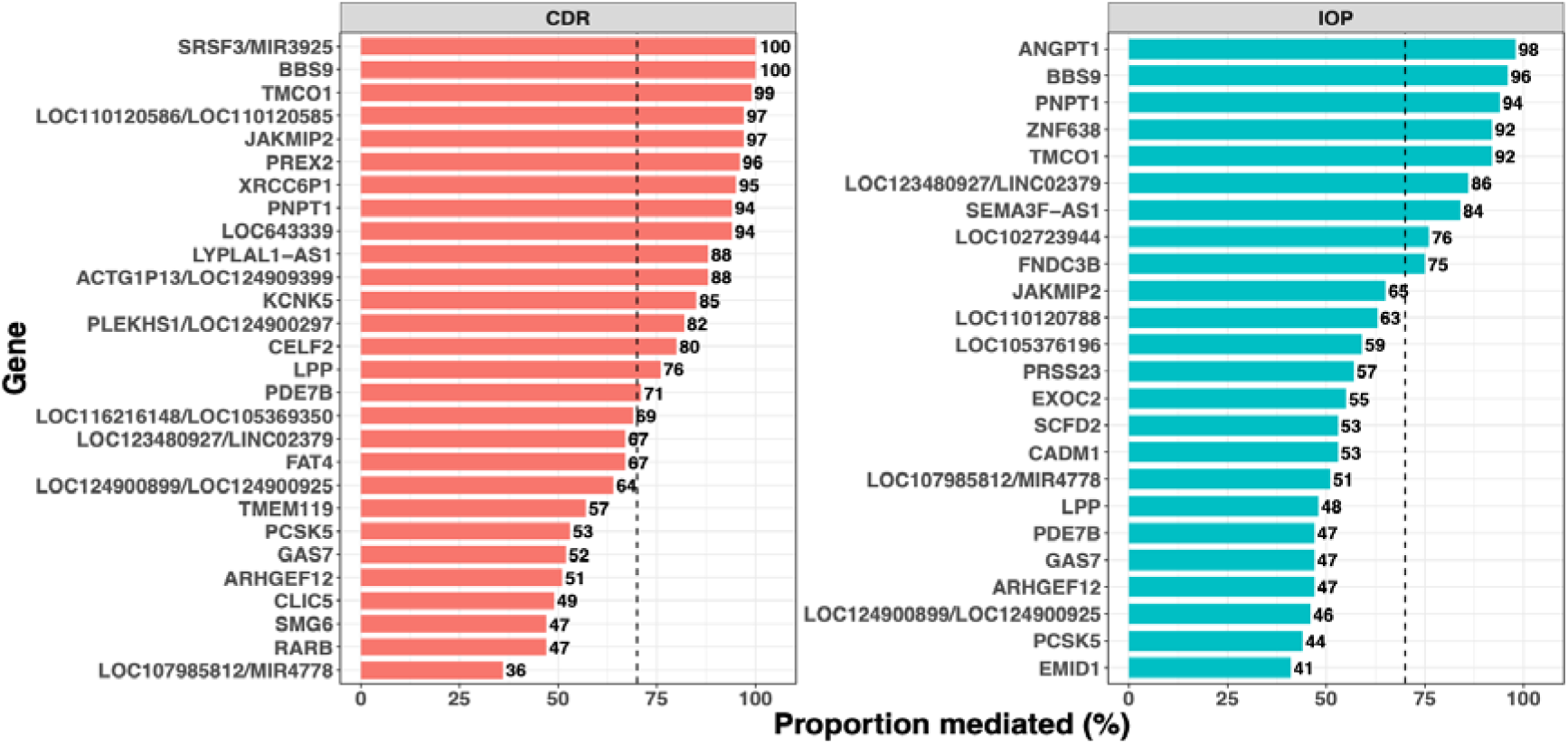
Estimated proportion of variant effects on POAG mediated through IOP and CDR. Variants were annotated to their nearest genes using Biofilter. For genes represented by multiple variants, the variant with the strongest POAG association was retained. The dashed line indicates the 70% threshold which separates partial from full mediation.

Overall, a variant mapping to *TMCO1* exhibited evidence of full mediation through both IOP (92%) and CDR (99%), suggesting that the effect of this locus on POAG may be largely mediated through both IOP and CDR. Consistent with our findings, *TMCO1* is a well-established POAG susceptibility locus that has previously been associated with both POAG and IOP.^41,42^ Similarly, a variant mapping to the *ARHGEF12* locus demonstrated partial mediation through both IOP (47%) and CDR (51%). *ARHGEF12* has also been identified as a POAG susceptibility gene, with previous studies suggesting that its effects on disease risk are mediated through both IOP and CDR.^4^Moreover, variants mapping to *GAS7* also showed similar patterns of partial mediation through IOP (47%) and CDR (52%).

On the other end, some loci exhibited endophenotype-specific patterns of mediation. For example, a variant (rs11556027) at the *PREX2* locus demonstrated 96% mediation through CDR. In contrast, variants at the *ANGPT1* locus exhibited up to 98% mediation through IOP. Consistent with previous studies, *ANGPT1* has been linked to IOP and POAG.^12,43^ In addition a locus mapping to the *FNDC3B* gene shows 73% proportion mediation through IOP; this gene has been previously associated with IOP.^43^

### Sensitivity Analysis of Existing Polygenic Risk Prediction Approaches

To benchmark the proposed endophenotype-informed PGS framework, we evaluated the performance of existing polygenic prediction approaches in both the POAAGG testing cohort (Table 4) and the PMBB validation cohort (Table 5). In the PMBB cohort, the baseline model (age, sex, and principal components) achieved an AUROC of 0.66. Adding a 568-SNP PGS constructed from the POAG GWAS produced only a modest improvement (AUROC = 0.68), and a 568-SNP PGS derived from MTAG summary statistics performed similarly (AUROC = 0.67), indicating limited predictive value from genetic information alone beyond baseline demographic and ancestry covariates. Genome-wide PGS constructed using the full POAG GWAS or full MTAG summary statistics performed comparably to the 568-SNP panels (AUROC = 0.69 for both), suggesting that expanding variant coverage from 568 SNPs to genome-wide scale provided minimal additional discriminative value in this cohort. In contrast, incorporating measured IOP or CDR substantially improved model performance regardless of whether a genetic score was included: models combining the 568-SNP PGS with IOP or CDR achieved an AUROC of 0.82, matching the performance of IOP- or CDR-only models without any genetic component (Models 2–3 vs. 8–9), indicating that the predictive gain was driven primarily by the endophenotype rather than the added genetic information. A broadly consistent pattern was observed in the POAAGG testing cohort, where models incorporating IOP or CDR outperformed genetic-only models. The near-perfect discrimination observed for the CDR-inclusive model likely reflects the inclusion of these endophenotypes in the POAAGG case definition, as described in the Methods. Taken together, these findings indicate that the observed improvement in predictive performance was primarily attributable to incorporation of the endophenotype, and that neither a curated nor a genome-wide polygenic score, on its own, approached the discriminative performance achieved by measured IOP or CDR.

**Table 4:**
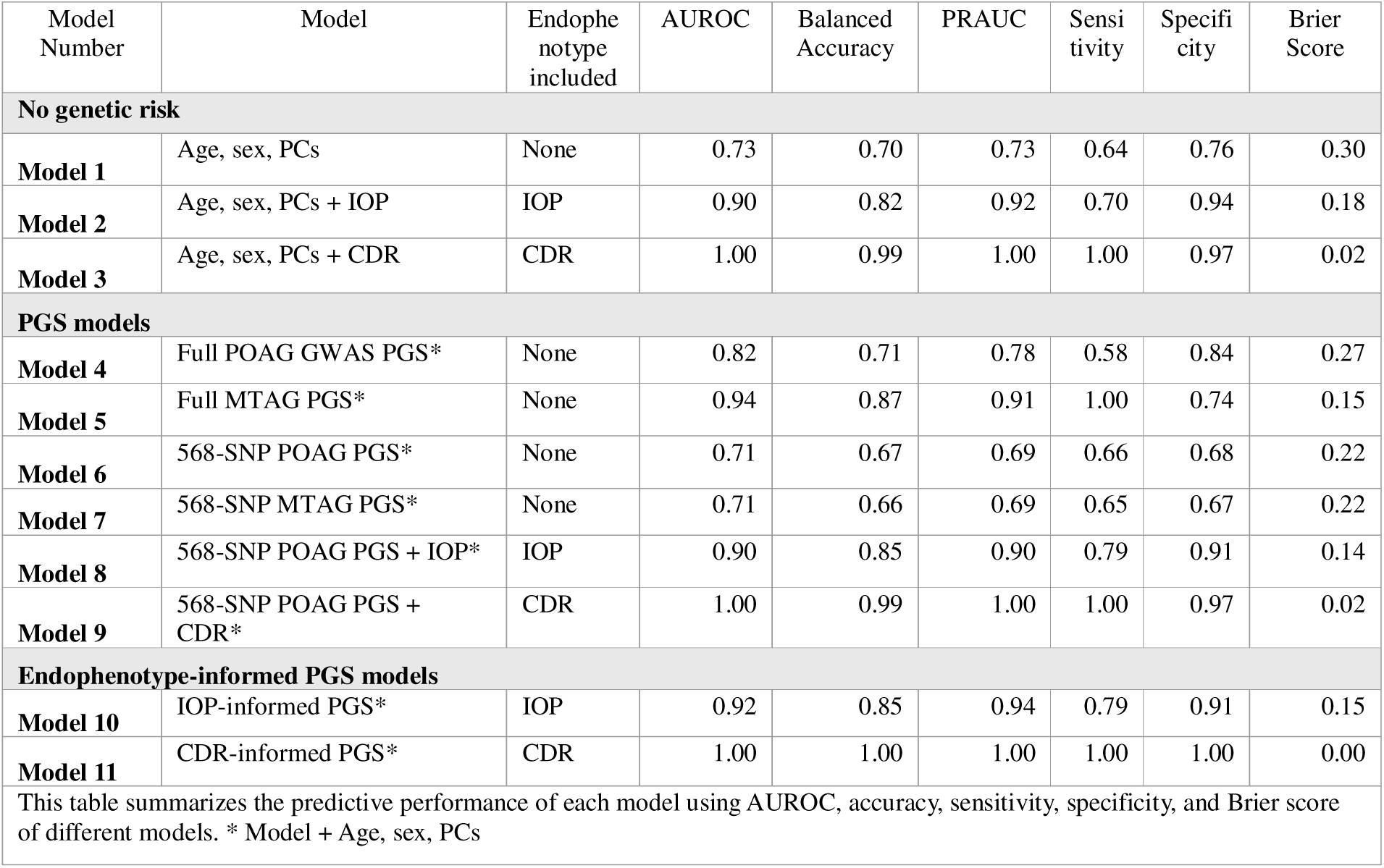
Performance metrics on POAAGG testing cohort comparing different models.

| Model Number | Model | Endophenotype included | AUROC | Balanced Accuracy | PRAUC | Sensitivity | Specificity | Brier Score |
| --- | --- | --- | --- | --- | --- | --- | --- | --- |
| **No genetic risk** |  |  |  |  |  |  |  |  |
| **Model 1** | Age, sex, PCs | None | 0.73 | 0.70 | 0.73 | 0.64 | 0.76 | 0.30 |
| **Model 2** | Age, sex, PCs + IOP | IOP | 0.90 | 0.82 | 0.92 | 0.70 | 0.94 | 0.18 |
| **Model 3** | Age, sex, PCs + CDR | CDR | 1.00 | 0.99 | 1.00 | 1.00 | 0.97 | 0.02 |
| **PGS models** |  |  |  |  |  |  |  |  |
| **Model 4** | Full POAG GWAS PGS* | None | 0.82 | 0.71 | 0.78 | 0.58 | 0.84 | 0.27 |
| **Model 5** | Full MTAG PGS* | None | 0.94 | 0.87 | 0.91 | 1.00 | 0.74 | 0.15 |
| **Model 6** | 568-SNP POAG PGS* | None | 0.71 | 0.67 | 0.69 | 0.66 | 0.68 | 0.22 |
| **Model 7** | 568-SNP MTAG PGS* | None | 0.71 | 0.66 | 0.69 | 0.65 | 0.67 | 0.22 |
| **Model 8** | 568-SNP POAG PGS + IOP* | IOP | 0.90 | 0.85 | 0.90 | 0.79 | 0.91 | 0.14 |
| **Model 9** | 568-SNP POAG PGS + CDR* | CDR | 1.00 | 0.99 | 1.00 | 1.00 | 0.97 | 0.02 |
| **Endophenotype-informed PGS models** |  |  |  |  |  |  |  |  |
| **Model 10** | IOP-informed PGS* | IOP | 0.92 | 0.85 | 0.94 | 0.79 | 0.91 | 0.15 |
| **Model 11** | CDR-informed PGS* | CDR | 1.00 | 1.00 | 1.00 | 1.00 | 1.00 | 0.00 |
| This table summarizes the predictive performance of each model using AUROC, accuracy, sensitivity, specificity, and Brier score of different models. * Model + Age, sex, PCs |  |  |  |  |  |  |  |  |

**Table 5:** Performance metrics on PMBB validation cohort comparing different models.

|  |  |  |  |  |  |  |  |  |
| --- | --- | --- | --- | --- | --- | --- | --- | --- |
| Model Number | Model | Endophenotype included | AUROC | Balanced Accuracy | PRAUC | Sensitivity | Specificity | Brier Score |
| **No genetic risk** |  |  |  |  |  |  |  |  |
| **Model 1** | Age, sex, PCs | None | 0.66 | 0.63 | 0.17 | 0.63 | 0.49 | 0.48 |
| **Model 2** | Age, sex, PCs + IOP | IOP | 0.82 | 0.72 | 0.55 | 0.70 | 0.74 | 0.26 |
| **Model 3** | Age, sex, PCs + CDR | CDR | 0.82 | 0.74 | 0.74 | 0.68 | 0.90 | 0.13 |
| **PGS models** |  |  |  |  |  |  |  |  |
| **Model 4** | Full POAG GWAS PGS* | None | 0.69 | 0.67 | 0.19 | 0.83 | 0.52 | 0.45 |
| **Model 5** | Full MTAG PGS* | None | 0.69 | 0.66 | 0.19 | 0.75 | 0.57 | 0.42 |
| **Model 6** | 568-SNP POAG PGS* | None | 0.68 | 0.61 | 0.17 | 0.83 | 0.40 | 0.56 |
| **Model 7** | 568-SNP MTAG PGS* | None | 0.67 | 0.63 | 0.17 | 0.80 | 0.46 | 0.51 |
| **Model 8** | 568-SNP POAG PGS + IOP* | IOP | 0.82 | 0.73 | 0.57 | 0.78 | 0.70 | 0.29 |
| **Model 9** | 568-SNP POAG PGS + CDR* | CDR | 0.82 | 0.76 | 0.47 | 0.62 | 0.88 | 0.13 |
| **Endophenotype-informed PGS models** |  |  |  |  |  |  |  |  |
| **Model 10** | IOP-informed PGS* | IOP | 0.82 | 0.75 | 0.39 | 0.65 | 0.84 | 0.18 |
| **Model 11** | CDR-informed PGS* | CDR | 0.82 | 0.74 | 0.52 | 0.73 | 0.76 | 0.24 |
| This table summarizes the predictive performance of each model using AUROC, accuracy, sensitivity, specificity, and Brier score of different models. * Model + Age, sex, PCs |  |  |  |  |  |  |  |  |

#### Predictive Performance of the Endophenotype-informed PGS

We next evaluated whether integrating endophenotype information directly into PGS construction could improve POAG risk prediction. Unlike conventional prediction models, which treat endophenotypes such as IOP or CDR as independent covariates, the endophenotype-informed PGS incorporates both the measured endophenotype value and mediator-specific SNP effect estimates into a single integrated risk score. Specifically, SNP effect estimates derived from the endophenotype GWAS and mediation analyses were combined according to Eq. (4), allowing the contribution of each genetic variant to vary as a function of the corresponding endophenotype. Two prediction models were evaluated: a CDR-informed PGS incorporating mediator-specific SNP effects and measured CDR values (model 10); and an IOP-informed PGS incorporating mediator-specific SNP effects and measured IOP values (model 11).

First, we evaluated the endophenotype-informed PGSs in a PGS testing cohort derived from an independent holdout subset of the POAAGG study. Compared with the baseline model, both endophenotype-informed PGSs demonstrated substantially improved predictive performance. The CDR-informed PGS achieved an AUROC of 1.00, whereas the IOP-informed PGS achieved an AUROC of 0.92, after adjustment for age, sex, and the first four principal components (Table 4). Complete model performance metrics, including AUROC, balanced accuracy, PRAUC, sensitivity, specificity, and Brier score, are summarized in Table 4. The near-perfect discrimination observed for the CDR-informed is consistent with the CDR-dependent case ascertainment in this cohort as described in the Methods.

To further assess the robustness and generalizability of these findings, we evaluated the endophenotype-informed PGSs in the independent PMBB validation cohort using the same prediction framework. In this cohort, the baseline model demonstrated modest discrimination for POAG (AUROC = 0.66). In contrast, both endophenotype-informed PGSs substantially improved predictive performance. The CDR-informed PGS achieved an AUROC of 0.82, while the IOP-informed PGS also achieved an AUROC of 0.82, after adjustment for age, sex, and the first four principal components. Complete model performance metrics, including AUROC, balanced accuracy, PRAUC, sensitivity, specificity, and Brier score, are presented in Table 5.

The improved performance observed in this independent validation cohort further supports the utility of incorporating measured endophenotype values into polygenic risk prediction. In addition to improving AUROC, the endophenotype-informed PGSs substantially increased PRAUC. Given the class imbalance in the validation cohort (177 cases and 1,486 controls), PRAUC provides a more informative measure of the models’ ability to identify true POAG cases. The IOP- and CDR-informed PGSs achieved PRAUC values of 0.52 and 0.39, respectively, compared with 0.17 for the baseline models. Considering that the expected PRAUC of a random classifier is approximately equal to the disease prevalence in the cohort (10.6%), the IOP- and CDR-informed PGSs demonstrated improvements over random classification. These results suggest that, decomposing genetic effects into endophenotype-mediated and direct components is valuable for identifying the specific pathways through which individual risk loci act, information not recoverable from previously established PGSs which provides an aggregated score.

#### Risk Stratification in the POAAGG Suspect Cohort

We next evaluated the endophenotype-informed PGS in a cohort of POAAGG suspects, divided into converters and non-converters. In this analysis, the IOP-informed PGS showed significantly higher separation between converters and non-converters (Figure 3), supporting its utility for risk stratification in individuals at elevated baseline risk. In contrast, the CDR-informed PGS did not show significant separation between converters and non-converters in this cohort.

**Figure 3.**
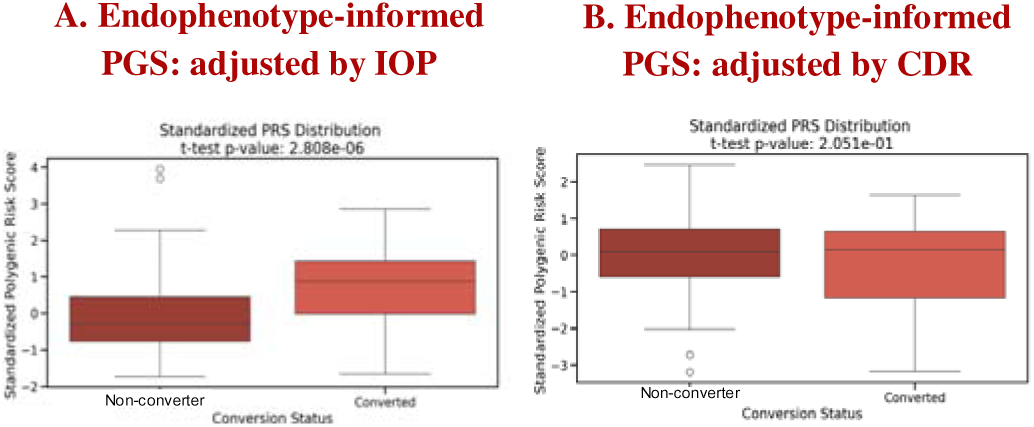
Endophenotype-informed PGS improved stratification of POAG progress among suspects: This figure compares the ability of IOP or CDR-informed PGSs to distinguish between POAG converters and non-converters suspects.

#### Endophenotype-informed PGS Captures Disease Heterogeneity Beyond Case-Control Status

We identified PMBB participants in the top 5% and bottom 5% of the PGS distribution with discordant risk level and disease status. The IOP-informed PGS identified 38 controls in the top 5% and 2 cases in the bottom 5%, while the CDR-informed PGS identified 41 controls in the top 5% and 3 cases in the bottom 5% (Supplementary Table S8).

Among the 41 controls identified by the CDR-informed PGS in the top 5% of genetic risk, at baseline visit, all were determined through chart review to be POAG suspects rather than true controls, indicating that they had been misclassified in the biobank phenotyping. Clinically, these high-risk individuals were characterized by structural vulnerability to POAG (higher mean CDR, borderline RNFL thickness), despite controlled IOP levels, with almost one-third converting from suspect to case over study duration. In the bottom 5% of risk of the CDR-informed PGS, two of the three POAG cases met the criteria for OHT, while the third was classified as a POAG suspect at baseline and subsequently converted to POAG during follow-up. These individuals exhibited elevated IOP_max yet had little structural or functional damage and slow progression rates. Overall, these results support that the CDR-informed PGS captures a continuum of susceptibility to structural glaucoma damage, with the ability to capture EHR misclassification among high-risk controls.

Among the 38 controls identified by the IOP-informed PGS in the top 5% of genetic risk, 53% were determined through chart review to be POAG suspects at baseline rather than true unaffected controls, indicating further misclassification within the biobank. Clinically, these individuals were characterized by consistently elevated IOP, yet they exhibited limited structural damage, slow progression rates, and low conversion rates (12%) during the follow-up period. Among the two POAG cases identified within the bottom 5% of genetic risk by the IOP-informed PGS, one had NTG and the other had OHT. Both individuals showed signs of structural and functional glaucomatous damage (mean MD=-6.7 dB), despite low and controlled IOP levels.

### DISCUSSION

In this study, we evaluated whether genetic risk prediction for primary open-angle glaucoma (POAG) could be improved and made more clinically interpretable by incorporating POAG-related endophenotypes. Using African ancestry cohorts from POAAGG and PMBB, we compared multiple sources of genetic risk information, including genome-wide POAG GWAS, MTAG-derived GWAS integrating POAG with IOP and CDR, selected SNP sets derived from these GWAS, and endophenotype-informed scores generated from SNP-level mediation analyses. We then evaluated these models in testing, external validation, longitudinal glaucoma-suspect, and manual chart-review analyses. Sensitivity analyses provide an important context for interpreting the prediction results. The 568-SNP PGS alone provided only modest improvement over baseline demographic and ancestry covariates. In contrast, incorporating clinically measured IOP or CDR substantially improved predictive performance across multiple metrics. Adding the 568-SNP PGS to models that already included these endophenotypes yielded little to no additional improvement in AUROC, suggesting that the predictive gain was driven primarily by the measured endophenotype itself rather than by added genetic information. These findings underscore the central role of routinely measured ocular traits in POAG risk stratification and highlight the need to interpret PGS-based models in relation to the clinical variables through which disease risk is expressed. This distinction is important for interpreting the endophenotype-informed PGS framework. POAG was modeled as a polygenic disease, but the aggregate PGS itself was not used to decompose genetic risk into direct and indirect effects. Effect decomposition was performed at the SNP level using counterfactual mediation analyses, and the resulting estimates were subsequently aggregated into PGS. Therefore, the primary contribution of the mediation framework is not simply improved discrimination beyond models that include measured IOP or CDR. Its value lies in linking inherited genetic risk to measurable ocular traits, enabling pathway-informed interpretation of how genetic susceptibility may be expressed clinically.

The mediation analyses provided examples of this interpretability. *ARHGEF12* showed partial mediation through both IOP (47%) and CDR (51%), consistent with its role in the RhoA/ROCK pathway. This gene activates RhoA, leading to a reduction in aqueous humor and resultant increase in IOP and CDR, particularly in individuals of African ancestry.^4^ *TMCO1* (transmembrane and coiled-coil domain 1) showed near-complete mediation through both IOP and CDR (92% and 99%, respectively). This finding aligns with experimental evidence linking this gene to trabecular meshwork dysfunction (RK/MAPK signaling pathway) and impaired aqueous humor outflow, leading to elevated IOP and supporting *TMCO1* as a potential therapeutic target for POAG. Although a direct mechanistic link between *TMCO1* and CDR has not yet been established, the near-complete mediation through CDR observed in our study may reflect the downstream consequences of sustained IOP elevation, whereby progressive optic nerve damage results in increased optic disc cupping.^42^ Another notable finding was that *ANGPT1* (angiopoietin-1) exhibited near complete (98%) mediation through IOP, consistent with its role in Schlemm’s canal development and the primary aqueous humor drainage pathway.^47,48^ In mouse models, loss of *ANGPT1* impairs aqueous humor outflow, resulting in elevated IOP. Furthermore, *ANGPT1* variants have also been identified in glaucoma patients, highlighting the clinical relevance of this pathway.^47,48^ These examples support the value of mediation analysis for connecting genetic associations to clinically measurable pathways, providing insight beyond conventional GWAS associations.

Our comparison of genome-wide and selected-SNP approaches has practical implications for clinical translation. Genome-wide PGS models, including MTAG-derived scores with approximately 1.1 million variants, provided only modest improvement over selected 568-SNP models in genetics-only prediction. This genome-wide approach modestly outperformed the 568-SNP PGS on genetics-only prediction (AUROC 0.69 vs. 0.68; PRAUC 0.19 vs. 0.17), but the improvement was small relative to the roughly 2,000-fold increase in variant number. This small gain should be weighed against the larger variant burden, lower transparency, and greater implementation complexity of genome-wide scores. A curated selected-SNP approach may offer advantages in interpretability, computational efficiency, and clinical communication, especially when discrimination is similar. This is particularly relevant given prior evidence from studies like in coronary heart disease showing that PGS with similar population-level performance can produce discordant individual-level risk estimates^49,50^. For POAG, where risk scores may influence screening intensity and longitudinal monitoring, interpretability and individual-level consistency are clinically important, not secondary to marginal gains in AUROC.

Building upon these findings, the endophenotype-informed PGS framework integrated measured IOP and CDR with SNP-level mediation estimates, rather than modeling endophenotypes as independent covariates. This approach achieved predictive performance comparable to conventional models that included measured IOP or CDR while substantially outperforming the 568-SNP PGS alone. Thus, the main advantage of the endophenotype-informed framework was not superior discrimination beyond models that already included the endophenotype, but a more interpretable representation of how inherited risk aligns with clinically measured disease-related traits. By embedding IOP- and CDR-related information into pathway-informed scores, this framework may support more clinically meaningful risk stratification for complex diseases such as POAG.

The longitudinal and chart-review analyses extended these findings beyond standard case-control prediction. In the POAG suspect cohort, the IOP-informed model better distinguished individuals who later converted to POAG from those who remained stable, suggesting that pressure-related endophenotype information may help identify individuals at higher risk before definitive diagnosis. In PMBB, manual chart review of participants with discordant PGS and EHR-derived disease status showed that some individuals labeled as controls had clinical features consistent with glaucoma suspect status. These findings suggest that models integrating genetic risk with measured endophenotypes may help identify EHR misclassification and clinically relevant heterogeneity in real-world health system data.

The CDR-informed analyses require particular caution. Because enlarged CDR is not specific to glaucoma, CDR-informed scores should not be interpreted as standalone evidence of glaucomatous damage. Their potential value is more appropriately framed as identifying individuals whose optic nerve structure may warrant closer clinical review or longitudinal follow-up. In contrast, IOP-informed models may be more relevant for identifying individuals with pressure-related risk before definitive diagnosis. These interpretations remain exploratory, particularly because the chart-review analyses involved small numbers of individuals at the extremes of the score distributions.

Several limitations should be considered. First, the modest sample limited the power for endophenotype GWAS where quantitative trait variation is more subtle. Second, the POAAGG testing cohort consisted of selected cases and controls, which may introduce selection bias or overfitting. Third, our downstream analyses focused exclusively on IOP and CDR, although other POAG-related endophenotypes may also contribute to disease risk. Fourth, because PMBB data were extracted from the EHR, measured endophenotypes may not always have preceded POAG diagnosis and may have been influenced by treatment, disease monitoring, or disease severity. This limits temporal and causal interpretation, particularly for mediation analyses in real-world EHR-derived data. Fifth, EHR-derived case-control labels are imperfect, as supported by the chart-review findings. Finally, all analyses were conducted in individuals of African ancestry. Although this focus addresses a major gap in glaucoma genetics, validation in additional ancestry groups will be necessary before broader clinical implementation.

Future work should evaluate multi-mediator all endophenotype-informed models that jointly incorporate multiple clinically relevant traits, rather than modeling IOP and CDR separately. Such models may better capture the interacting pathways that underlie POAG risk. Larger longitudinal cohorts will also be needed to assess calibration, clinical thresholds, and decision-curve performance, and to determine whether these scores improve screening, referral, or monitoring decisions in real-world care.

In summary, endophenotype-informed GWAS and PGS analyses provide a clinically interpretable framework for POAG genetic risk with routinely measured ocular traits. Although measured IOP and CDR accounted for much of the improvement in predictive performance, SNP-level mediation analyses helped organize genetic risk around clinically meaningful pathways. These findings support integrating inherited susceptibility with ocular endophenotypes as a promising strategy for earlier, pathway-informed POAG risk stratification in populations disproportionately affected by this blinding disease.

## Data Availability

Genotype and phenotype data for POAAGG subjects is available on dbGAP (accession phs001312).

https://github.com/Setia-Verma-Lab/POAG_PSB_paper_supplementary_tables

## ACKNOWLEDGEMENTS

The Primary Open-Angle African Ancestry Glaucoma Genetics (POAAGG) study is supported by the National Eye Institute, Bethesda, Maryland (grant #1R01EY023557) and the Vision Research Core Grant (P30EY001583-51). Funds also come from the F.M. Kirby Foundation, Research to Prevent Blindness, The UPenn Hospital Board of Women Visitors, and The Paul and Evanina Bell Mackall Foundation Trust. Support also came from Regeneron Genetics Center, the Ophthalmology Department at the Perelman School of Medicine, the Genetics Department at the Perelman School of Medicine, and the VA Hospital in Philadelphia, PA. This work was also supported by NIH grants R01DK134575 (PI: Dr. Vujkovic) and R01HD110567 (PI: Dr. Setia-Verma), which funded research underlying the development of the mediator-effect PRS model. We greatly appreciate the work of the Clinical Research Coordinators in the POAAGG study, who made it possible to enroll more than 10,200 individuals of African ancestry in Philadelphia. The funding sources had no role in the design and conduct of the study; collection, management, analysis, and interpretation of the data; preparation, review, or approval of the manuscript; or the decision to submit the manuscript for publication.

## AUTHOR CONTRIBUTIONS

Conceptualization: Aude Benigne Ikuzwe Sindikubwabo, Shefali Setia-Verma

Data Curation: Aude Benigne Ikuzwe Sindikubwabo, Yan Zhu, Yuki Bradford, Colleen Moorse, Anurag Verma

Formal analysis: Aude Benigne Ikuzwe Sindikubwabo, Lannawill Caruth, Yuki Bradford

Funding Acquisition: Joan M. O’Brien, Shefali Setia-Verma, Marijana Vujković

Investigation: Yan Zhu, Rebecca Salowe, Roy Lee, Leila Ghaffari, Mina Halimitabrizi

Methodology: Marijana Vujković, Shefali Setia-Verma, Aude Benigne Ikuzwe Sindikubwabo

Project administration: Rebecca Salowe, Shefali Setia-Verma, Joan M. O’Brien, Marylyn D. Ritchie, Ahmara Gibbons Ross

Resources: Shefali Setia-Verma, Joan M. O’Brien, Marylyn D. Ritchie

Software: Aude Benigne Ikuzwe, Lannawill Caruth, Yuki Bradford

Supervision: Marylyn D. Ritchie, Shefali Setia-Verma, Joan M. O’Brien, Ahmara Gibbons Ross

Writing – Original Draft: Aude Benigne Ikuzwe Sindikubwabo, Shefali Setia-Verma

Writing – Review & Editing: all co-authors

## DATA AVAILABILITY

- The de-identified genotype data used in the POAAGG study are available through dbGAP accession number phs001312. Due to POAAGG subject privacy concerns and IRB regulations, identifiable data cannot be deposited in a public repository. However, de-identified datasets used for model training and evaluation are available from the lead contact upon request and pending data use agreement.
- Supplementary materials are available in the attached spreadsheet.

## DECLARATION OF GENERATIVE AI AND AI-ASSISTED TECHNOLOGIES IN THE MANUSCRIPT PREPARATION PROCESS

During the preparation of this manuscript, the authors used ChatGPT and Claude to assist with language editing and refinement. The author(s) reviewed and edited the output as needed and take full responsibility for the content of the published article.

## REFERENCES

1. Weinreb RN, Khaw PT. Primary open-angle glaucoma. The Lancet. 2004 May;363(9422):1711–20. doi:10.1016/S0140-6736(04)16257-0

2. Allison K, Patel D, Alabi O. Epidemiology of Glaucoma: The Past, Present, and Predictions for the Future. Cureus. 2020 Nov 24;12(11):e11686. doi:10.7759/cureus.11686 PubMed PMID: 33391921; PubMed Central PMCID: PMC7769798.

3. Zhang N, Wang J, Li Y, Jiang B. Prevalence of primary open angle glaucoma in the last 20 years: a meta-analysis and systematic review. Sci Rep. 2021 Jul 2;11(1):13762. doi:10.1038/s41598-021-92971-w PubMed PMID: 34215769; PubMed Central PMCID: PMC8253788.

4. Verma SS, Gudiseva HV, Chavali VRM, Salowe RJ, Bradford Y, Guare L, et al. A multi-cohort genome-wide association study in African ancestry individuals reveals risk loci for primary open-angle glaucoma. Cell. 2024 Jan 18;187(2):464–480.e10. doi:10.1016/j.cell.2023.12.006 PubMed PMID: 38242088; PubMed Central PMCID: PMC11844349.

5. Tham YC, Li X, Wong TY, Quigley HA, Aung T, Cheng CY. Global prevalence of glaucoma and projections of glaucoma burden through 2040: a systematic review and meta-analysis. Ophthalmology. 2014 Nov;121(11):2081–90. doi:10.1016/j.ophtha.2014.05.013 PubMed PMID: 24974815.

6. Asefa NG, Neustaeter A, Jansonius NM, Snieder H. Heritability of glaucoma and glaucoma-related endophenotypes: Systematic review and meta-analysis. Surv Ophthalmol. 2019;64(6):835–51. doi:10.1016/j.survophthal.2019.06.002 PubMed PMID: 31229521.

7. Springelkamp H, Iglesias AI, Mishra A, Höhn R, Wojciechowski R, Khawaja AP, et al. New insights into the genetics of primary open-angle glaucoma based on meta-analyses of intraocular pressure and optic disc characteristics. Hum Mol Genet. 2017 Jan 15;26(2):438–53. doi:10.1093/hmg/ddw399 PubMed PMID: 28073927; PubMed Central PMCID: PMC5968632.

8. Gharahkhani P, Jorgenson E, Hysi P, Khawaja AP, Pendergrass S, Han X, et al. Genome-wide meta-analysis identifies 127 open-angle glaucoma loci with consistent effect across ancestries. Nat Commun. 2021 Feb 24;12(1):1258. doi:10.1038/s41467-020-20851-4 PubMed PMID: 33627673; PubMed Central PMCID: PMC7904932.

9. Fan BJ, Wang DY, Pasquale LR, Haines JL, Wiggs JL. Genetic variants associated with optic nerve vertical cup-to-disc ratio are risk factors for primary open angle glaucoma in a US Caucasian population. Invest Ophthalmol Vis Sci. 2011 Mar 28;52(3):1788–92. doi:10.1167/iovs.10-6339 PubMed PMID: 21398277; PubMed Central PMCID: PMC3101676.

10. Solovieff N, Cotsapas C, Lee PH, Purcell SM, Smoller JW. Pleiotropy in complex traits: challenges and strategies. Nat Rev Genet. 2013 Jul;14(7):483–95. doi:10.1038/nrg3461 PubMed PMID: 23752797; PubMed Central PMCID: PMC4104202.

11. Craig JE, Han X, Qassim A, Hassall M, Cooke Bailey JN, Kinzy TG, et al. Multitrait analysis of glaucoma identifies new risk loci and enables polygenic prediction of disease susceptibility and progression. Nat Genet. 2020 Feb;52(2):160–6. doi:10.1038/s41588-019-0556-y PubMed PMID: 31959993; PubMed Central PMCID: PMC8056672.

12. Han X, Gharahkhani P, Hamel AR, Ong JS, Rentería ME, Mehta P, et al. Large-scale multitrait genome-wide association analyses identify hundreds of glaucoma risk loci. Nat Genet. 2023 Jul;55(7):1116–25. doi:10.1038/s41588-023-01428-5 PubMed PMID: 37386247; PubMed Central PMCID: PMC10335935.

13. Lo Faro V, Bhattacharya A, Zhou W, Zhou D, Wang Y, Läll K, et al. Novel ancestry-specific primary open-angle glaucoma loci and shared biology with vascular mechanisms and cell proliferation. Cell Rep Med. 2024 Feb 20;5(2):101430. doi:10.1016/j.xcrm.2024.101430 PubMed PMID: 38382466; PubMed Central PMCID: PMC10897632.

14. Chang-Wolf JM, Kinzy TG, Driessen SJ, Cruz LA, Iyengar SK, Peachey NS, et al. Performance of Polygenic Risk Scores for Primary Open-Angle Glaucoma in Populations of African Descent. JAMA Ophthalmol. 2025 Jan 1;143(1):7–14. doi:10.1001/jamaophthalmol.2024.4784 PubMed PMID: 39541127; PubMed Central PMCID: PMC11565374.

15. Marijana V, David E. Kaplan4,1,*, Jonas Ghouse5,6,7, Bao-Li Loza8, Joseph Brancale9,10, Adam Lewis11, David Y. Zhang12, Michael G. Levin13,14,1, Olivia J. Veatch15, Josephine P. Johnson2, Carolin V. Schneider16,17,2, Anurag Verma2,1, Kirk J. Wangensteen18, Eleonora Scorletti2, Dipender Gill19, Chigoziri Konkwo9, Alexis M. Garófalo2, Lindsay A. Guare20, Tae-Wi Schwantes-An21,22, Marco V. Abreu21,22, Kyung Min Lee23, Abraham Shaked24, Kim M. Olthoff24, Maarouf A. Hoteit4, Elizabeth K. Speliotes25,26, Yanhua Chen25, Antonino Oliveri25, Lishi Yin25, Luca VC. Valenti27,28, Francesco Malvestiti27, Daniele Marchelli27, Lorenzo Miano27,28, Quentin M. Anstee29,30, Ann K. Daly29, Heather J. Cordell31, Rebecca Darlay31, Niek Verweij32, George Hindy32, Adam Locke32, Kentaro Matsuura33, Sumeet K. Asrani34, James F. Trotter34, Giuliano Testa34, Luca A. Lotta32, Marcus B. Jones32, Daniel R. Dochtermann35, Trina M. Norden-Krichmar22,36, Craig C. Teerlink23,37, Poornima Devineni35, Saiju Pyarajan35,38, Daniel J. Rader2,12, Yasuhito Tanaka39, Benjamin F. Voight1,12,40, Silvia Vilarinho9,10, Lisa A. Bastarache11, Stefan Stender41,42, Philip S. Tsao43,44, Penn Medicine Biobank, Copenhagen Hospital Biobank and Danish Blood and Danish Blood Donor Study, BioVU Biobank, Michigan Genomics Initiative, Regeneron Genetics Center, Indiana Biobank, AllOfUs Biobank, Milano Biobank, LITMUS Consortium, VA Million Veteran Program, Timothy R. Morgan22,45, Julie A. Lynch23,37, Kyong-Mi Chang4,1. Germline Variants Influence Chronic Liver Disease Progression through Distinct Pathways.

16. Salowe RJ, Lee R, Zenebe-Gete S, Vaughn M, Gudiseva HV, Pistilli M, et al. Recruitment strategies and lessons learned from a large genetic study of African Americans. PLOS Glob Public Health. 2022;2(8):e0000416. doi:10.1371/journal.pgph.0000416 PubMed PMID: 36743904; PubMed Central PMCID: PMC9897316.

17. Charlson ES, Sankar PS, Miller-Ellis E, Regina M, Fertig R, Salinas J, et al. The primary open-angle african american glaucoma genetics study: baseline demographics. Ophthalmology. 2015 Apr;122(4):711–20. doi:10.1016/j.ophtha.2014.11.015 PubMed PMID: 25576993; PubMed Central PMCID: PMC4372490.

18. van der Valk R, Webers CAB, Schouten JSAG, Zeegers MP, Hendrikse F, Prins MH. Intraocular pressure-lowering effects of all commonly used glaucoma drugs: a meta-analysis of randomized clinical trials. Ophthalmology. 2005 Jul;112(7):1177–85. doi:10.1016/j.ophtha.2005.01.042 PubMed PMID: 15921747.

19. Deubel C, Böhringer D, Anton A, Reinhard T, Lübke J. Long-term follow-up of intraocular pressure and pressure-lowering medication in patients following Excimer laser trabeculotomy. Graefes Arch Clin Exp Ophthalmol. 2021 Apr;259(4):957–62. doi:10.1007/s00417-020-05029-4 PubMed PMID: 33289863; PubMed Central PMCID: PMC8016798.

20. Harris PA, Taylor R, Thielke R, Payne J, Gonzalez N, Conde JG. Research electronic data capture (REDCap)--a metadata-driven methodology and workflow process for providing translational research informatics support. J Biomed Inform. 2009 Apr;42(2):377–81. doi:10.1016/j.jbi.2008.08.010 PubMed PMID: 18929686; PubMed Central PMCID: PMC2700030.

21. Harris PA, Taylor R, Minor BL, Elliott V, Fernandez M, O’Neal L, et al. The REDCap consortium: Building an international community of software platform partners. J Biomed Inform. 2019 Jul;95:103208. doi:10.1016/j.jbi.2019.103208 PubMed PMID: 31078660; PubMed Central PMCID: PMC7254481.

22. Zhao AT, Di Rosa I, Salowe R, Jin F, Lee R, Aibo MAI, et al. Predictors of Glaucoma Conversion in an African Ancestry Cohort: A Longitudinal Study. Am J Ophthalmol. 2025 Dec;280:120–9. doi:10.1016/j.ajo.2025.07.035 PubMed PMID: 40782885; PubMed Central PMCID: PMC12969997.

23. Verma A, Damrauer SM, Naseer N, Weaver J, Kripke CM, Guare L, et al. The Penn Medicine BioBank: Towards a Genomics-Enabled Learning Healthcare System to Accelerate Precision Medicine in a Diverse Population. J Pers Med. 2022 Nov 29;12(12):1974. doi:10.3390/jpm12121974 PubMed PMID: 36556195; PubMed Central PMCID: PMC9785650.

24. Das S, Forer L, Schönherr S, Sidore C, Locke AE, Kwong A, et al. Next-generation genotype imputation service and methods. Nat Genet. 2016 Oct;48(10):1284–7. doi:10.1038/ng.3656 PubMed PMID: 27571263; PubMed Central PMCID: PMC5157836.

25. Galinsky KJ, Loh PR, Mallick S, Patterson NJ, Price AL. Population Structure of UK Biobank and Ancient Eurasians Reveals Adaptation at Genes Influencing Blood Pressure. Am J Hum Genet. 2016 Nov 3;99(5):1130–9. doi:10.1016/j.ajhg.2016.09.014 PubMed PMID: 27773431; PubMed Central PMCID: PMC5097941.

26. Galinsky KJ, Bhatia G, Loh PR, Georgiev S, Mukherjee S, Patterson NJ, et al. Fast Principal-Component Analysis Reveals Convergent Evolution of ADH1B in Europe and East Asia. Am J Hum Genet. 2016 Mar 3;98(3):456–72. doi:10.1016/j.ajhg.2015.12.022 PubMed PMID: 26924531; PubMed Central PMCID: PMC4827102.

27. Zhou W, Nielsen JB, Fritsche LG, Dey R, Gabrielsen ME, Wolford BN, et al. Efficiently controlling for case-control imbalance and sample relatedness in large-scale genetic association studies. Nat Genet. 2018 Sep;50(9):1335–41. doi:10.1038/s41588-018-0184-y PubMed PMID: 30104761; PubMed Central PMCID: PMC6119127.

28. Turley P, Walters RK, Maghzian O, Okbay A, Lee JJ, Fontana MA, et al. Multi-trait analysis of genome-wide association summary statistics using MTAG. Nat Genet. 2018 Feb;50(2):229–37. doi:10.1038/s41588-017-0009-4 PubMed PMID: 29292387; PubMed Central PMCID: PMC5805593.

29. Chang CC, Chow CC, Tellier LC, Vattikuti S, Purcell SM, Lee JJ. Second-generation PLINK: rising to the challenge of larger and richer datasets. Gigascience. 2015;4:7. doi:10.1186/s13742-015-0047-8 PubMed PMID: 25722852; PubMed Central PMCID: PMC4342193.

30. Sobel ME. Asymptotic Confidence Intervals for Indirect Effects in Structural Equation Models. Sociological Methodology. 1982;13:290. doi:10.2307/270723

31. Vanderweele TJ, Vansteelandt S, Robins JM. Effect decomposition in the presence of an exposure-induced mediator-outcome confounder. Epidemiology. 2014 Mar;25(2):300–6. doi:10.1097/EDE.0000000000000034 PubMed PMID: 24487213; PubMed Central PMCID: PMC4214081.

32. Wald A. Tests of statistical hypotheses concerning several parameters when the number of observations is large. Trans Amer Math Soc. 1943;54(3):426–82. doi:10.1090/S0002-9947-1943-0012401-3

33. Ruan Y, Lin YF, Feng YCA, Chen CY, Lam M, Guo Z, et al. Improving polygenic prediction in ancestrally diverse populations. Nat Genet. 2022 May;54(5):573–80. doi:10.1038/s41588-022-01054-7 PubMed PMID: 35513724; PubMed Central PMCID: PMC9117455.

34. Robins JM, Greenland S. Identifiability and exchangeability for direct and indirect effects. Epidemiology. 1992 Mar;3(2):143–55. doi:10.1097/00001648-199203000-00013 PubMed PMID: 1576220.

35. MacKinnon DP, Lockwood CM, Hoffman JM, West SG, Sheets V. A comparison of methods to test mediation and other intervening variable effects. Psychol Methods. 2002 Mar;7(1):83–104. doi:10.1037/1082-989x.7.1.83 PubMed PMID: 11928892; PubMed Central PMCID: PMC2819363.

36. Valeri L, Vanderweele TJ. Mediation analysis allowing for exposure-mediator interactions and causal interpretation: theoretical assumptions and implementation with SAS and SPSS macros. Psychol Methods. 2013 Jun;18(2):137–50. doi:10.1037/a0031034 PubMed PMID: 23379553; PubMed Central PMCID: PMC3659198.

37. Park SH, Lee JS, Heo H, Suh YW, Kim SH, Lim KH, et al. A nationwide population-based study of low vision and blindness in South Korea. Invest Ophthalmol Vis Sci. 2014 Dec 18;56(1):484–93. doi:10.1167/iovs.14-14909 PubMed PMID: 25525165.

38. Kass MA, Heuer DK, Higginbotham EJ, Johnson CA, Keltner JL, Miller JP, et al. The Ocular Hypertension Treatment Study: a randomized trial determines that topical ocular hypotensive medication delays or prevents the onset of primary open-angle glaucoma. Arch Ophthalmol. 2002 Jun;120(6):701–13; discussion 829-830. doi:10.1001/archopht.120.6.701 PubMed PMID: 12049574.

39. Salowe RJ, Chen Y, Zenebe-Gete S, Lee R, Gudiseva HV, Di Rosa I, et al. Risk factors for structural and functional progression of primary open-angle glaucoma in an African ancestry cohort. BMJ Open Ophthalmol. 2023 Mar;8(1):e001120. doi:10.1136/bmjophth-2022-001120 PubMed PMID: 37278425; PubMed Central PMCID: PMC9990679.

40. Wang Z, Zhou H, Wang F, Huang H. Exploration of potential drug targets for Glaucoma by plasma proteome screening. J Proteomics. 2025 Jan 6;310:105324. doi:10.1016/j.jprot.2024.105324 PubMed PMID: 39342991.

41. Burdon KP, Macgregor S, Hewitt AW, Sharma S, Chidlow G, Mills RA, et al. Genome-wide association study identifies susceptibility loci for open angle glaucoma at TMCO1 and CDKN2B-AS1. Nat Genet. 2011 Jun;43(6):574–8. doi:10.1038/ng.824 PubMed PMID: 21532571.

42. Zhang Y, Han R, Xu S, Shen B, Yu H, Chen J, et al. TMCO1 promotes ferroptosis and ECM deposition in glaucomatous trabecular meshwork via ERK1/2 signaling. Biochim Biophys Acta Mol Basis Dis. 2025 Jan;1871(1):167530. doi:10.1016/j.bbadis.2024.167530 PubMed PMID: 39343416.

43. Choquet H, Thai KK, Yin J, Hoffmann TJ, Kvale MN, Banda Y, et al. A large multi-ethnic genome-wide association study identifies novel genetic loci for intraocular pressure. Nat Commun. 2017 Dec 13;8(1):2108. doi:10.1038/s41467-017-01913-6 PubMed PMID: 29235454; PubMed Central PMCID: PMC5727399.

44. George R, Panda S, Vijaya L. Blindness in glaucoma: primary open-angle glaucoma versus primary angle-closure glaucoma-a meta-analysis. Eye (Lond). 2022 Nov;36(11):2099–105. doi:10.1038/s41433-021-01802-9 PubMed PMID: 34645961; PubMed Central PMCID: PMC9582001.

45. Doozandeh A, Yazdani S, Pakravan M, Ghasemi Z, Hassanpour K, Hatami M, et al. Risk of Missed Diagnosis of Primary Open-Angle Glaucoma by Eye Care Providers. J Curr Ophthalmol. 2022;34(4):404–8. doi:10.4103/joco.joco_296_22 PubMed PMID: 37180528; PubMed Central PMCID: PMC10170989.

46. de Vries VA, Hanyuda A, Vergroesen JE, Do R, Friedman DS, Kraft P, et al. The Clinical Usefulness of a Glaucoma Polygenic Risk Score in 4 Population-Based European Ancestry Cohorts. Ophthalmology. 2025 Feb;132(2):228–37. doi:10.1016/j.ophtha.2024.08.005 PubMed PMID: 39128550; PubMed Central PMCID: PMC12204775.

47. Thomson BR, Heinen S, Jeansson M, Ghosh AK, Fatima A, Sung HK, et al. A lymphatic defect causes ocular hypertension and glaucoma in mice. J Clin Invest. 2014 Oct;124(10):4320–4. doi:10.1172/JCI77162 PubMed PMID: 25202984; PubMed Central PMCID: PMC4191022.

48. Thomson BR, Grannonico M, Liu F, Liu M, Mendapara P, Xu Y, et al. Angiopoietin-1 Knockout Mice as a Genetic Model of Open-Angle Glaucoma. Transl Vis Sci Technol. 2020 Mar;9(4):16. doi:10.1167/tvst.9.4.16 PubMed PMID: 32818103; PubMed Central PMCID: PMC7396191.

49. Chen GB, Lee SH, Montgomery GW, Wray NR, Visscher PM, Gearry RB, et al. Performance of risk prediction for inflammatory bowel disease based on genotyping platform and genomic risk score method. BMC Med Genet. 2017 Aug 29;18(1):94. doi:10.1186/s12881-017-0451-2 PubMed PMID: 28851283; PubMed Central PMCID: PMC5576242.

50. Ponelle-Chachuat F, Lepage M, Viala S, Kelleci M, Floch EL, Dandine-Roulland C, et al. Development and validation of a next-generation sequencing-based method for calculating the breast cancer polygenic risk score PRS313. Breast. 2025 Dec;84:104580. doi:10.1016/j.breast.2025.104580 PubMed PMID: 40974665; PubMed Central PMCID: PMC12481083.

